# Rationing and triage of scarce, lifesaving therapy in the context of the COVID-19 pandemic - a cross-sectional, social media-driven, scenario-based online query of societal attitudes

**DOI:** 10.1101/2020.07.28.20163360

**Authors:** Oliver J. Muensterer, Emilio A. Gianicolo, Norbert W. Paul

**Author notes:** Correspondence to: Oliver J. Muensterer, MD, PhD, Department of Pediatric Surgery, University Medical Center of the Johannes Gutenberg University Mainz, Langenbeckstrasse 1, 55131 Mainz, Germany. This work is part of the masterthesis of OM in medical ethics at the Institute for History, Theory and Ethics of Medicine at the Johannes Gutenberg University Mainz (under supervision of Norbert W. Paul).

## Abstract

**Background:** The recent COVID-19 pandemic made us aware that medical resources are limited. When demand for essential resources surpasses availability, difficult triaging decisions are necessary. While algorithms exist, almost nothing is known on societal attitudes regarding triage criteria.

**Methods:** A cross-sectional survey was conducted via social media channels to query a broad sample of society. Participants were asked to make triage decisions in case-based vignettes, exploring a variety of factors. They also were asked to assess how sure they were about their decisions, and how such decisions should be reached.

**Results:** The survey was completed in full by 1626 participants in April 2020. Median age was 39 years (range 12-80 years), 984 (61%) were female. Patient prognosis, responsibility towards others, the absence of behavior-induced co-morbidities, and younger age were rated the most important triage criteria, while participants found that insurance status, social status, and nationality should not play a substantial role. Ethics-committees and point systems were regarded potentially helpful for triage decision-making, while decisions based on order of presentation (first-come first-serve) or on a legal basis were viewed critically. Participants were least sure about their decision when dealing with age or behavior-induced co-morbidities. Overall, women were surer about their decisions than men, participants of Christian faith were also more secure about their decision than atheists-agnostics.

**Conclusions:** This study uses social media to generate insight into public opinion and attitudes regarding triage criteria and modalities. These findings may be helpful for the development of future medical triage algorithms.

## INTRODUCTION

Even in highly developed nations, the COVID-19 pandemic has shown how medical resources can become limited when an overwhelming need arises. The discrepancy between supply and demand in southern European medical centers, and later on in some metropolitan areas of the United States, reached dimensions previously unimaginable.[1] Some of the most strikingly extreme circumstances were the sudden need to ration lifesaving medical therapies, in particular intensive care beds and ventilators, including advanced technology such as extracorporal membrane oxygenation (ECMO).[2]

Italy was the first country in which, due to the vast number of affected patients with respiratory distress, medical personnel was required to make ad-hoc decisions regarding which patient would receive a ventilator or not.[3] This gut-wrenching task mostly fell on physicians working in the forefront, who were already overwhelmed and at their limits. The precarious, highly dynamic situation left them practically no opportunity to prepare emotionally, morally, or functionally for the resulting challenges.

Hence, the discourse focused on triage algorithms to be applied for the distribution of scarce therapeutic resources, mostly in an operational mode. Such practical guidelines have long been established in military and disaster.[4] Military algorithms, however, focus mainly on maintaining an effective army of soldiers. They do not lend themselves for simple translation into the civilian environment. After the September 11th attacks in 2001, algorithms with an emphasis on patient prognosis as the lead criterion were developed.[5] In accordance, with the principle of beneficence (acting with the best interest of the other), and with the utilitarian principle of conferring the most benefit to the greatest number of individuals, civilian triage systems correspondingly tend to favor patients with a better prognosis.

At times, triaging purely by prognosis seems inappropriate. Particularly the immanent conflict between urgency and the prospect of successful outcome, along with the need for a timely decision, poses an inextricable dilemma for medical practitioners in the trenches. Moral intuitions that overshadow rational decisions can complicate matters even further. For example, should a 30-year-old mother of 4 with a chronic medical condition receive priority in receiving a scarce, life-saving therapy over an otherwise healthy, single 65 year old? In reality, decisions are based on a multidimensional array of factors. These may include responsibility for others (as in this example), age, expected quality of life after convalescence, or sacrifice for others (for example, in the case of medical personnel). The weighting of these different factors is strongly dependent upon the moral norms that result from the discourse within a particular society. While experts and opinion leaders[6] have heavily driven the narrative, for example labelling triage according to age as “radical-utilitarian discrimination of the elderly”,[7] there is very little known about the opinions and attitudes of the general public. Cross sectional studies in this regard are lacking, but would be useful to guide the implementation of acceptable future triage algorithms.

These deficits have been addressed in several recent publications,[8] particularly in the context of the COVID-19 pandemic.[9] We were interested in surveying the opinions on concrete criteria that should play a role in the rationing of life-saving measures in a cross-sectional sample of the general public. Since contemporary public opinion has been shaped increasingly by social media, we decided to launch this empiric study of the societal attitudes through social media channels. After the COVID-19-pandemic had hit some European countries so heavily, conducting this study during the ongoing crisis was embedded in the ethically founded overall concept of preparedness, which might have contributed to the relatively controlled course of the pandemic in Germany with relatively low mortality rates. To our knowledge, a similarly-designed study has so far not been performed or published.

## METHODS

In April, 2020, an online survey (LimeSurvey GmbH, Hamburg) was generated in German language and propagated broadly via different social media channels (Internet webpages of our institution, medical ethics listserver, pediatric surgery forum, institutional Facebook account, Instagram account, SurveyCircle website, private contacts). In order to snowball the distributive effects, participants were expressively asked to resend the link to their friends and contacts.

Regarding content, the online survey (appendix A) included 10 imaginary scenarios, in which two critically-ill patients required mechanical ventilation, but only one ventilator was available. The fictitious patient pairs differed in one principal characteristic [1. age, 2. insurance status, 3. fateful, non-behavior-associated co-morbidities (such as other preexisting conditions), 4. behavior-associated co-morbidities (resulting from behavior, such as smoking or drinking alcohol), 5. working in a healthcare profession, 6. asylum status/nationality, 7. social status, 8. prognosis/chance of therapeutic success, 9. explicit consent for the intensive therapy, 10. responsibility for others]. The participants were asked who they would provide the ventilator to (patient A or patient B). Another option was to draw lots for an equal chance to receive the therapy. Subsequently, participants were asked to mark how sure they were of their decision on a scale from 0 to 100. They were also asked to mark how important in their opinion the particular criterion should be for the distribution of lifesaving therapies on a scale from 0 to 100.

After the imaginary case-vignettes, the participants were asked about their opinion on the modalities that should be employed to make triage decisions (a. drawing lots [randomization], b. chronologic distribution [“first come, first serve”], c. legal criteria [laws that specify how a physician should decide in a particular circumstance], d. ethics committees [on-site counselling of the physicians by an interdisciplinary team], e. decision committees [the decision is made by an independent team remote from the treatment location], f. a point-system similar to what is used for organ transplantation. Finally, the participants were queried on their age, sex, religious beliefs, and whether they were working in a medical profession.

Data were stored anonymously. The characteristics of our sample such as mean age, gender, religion and proportion of health care professional were compared to those of the general population (Statistisches Bundesamt[10]). Results of the scale values from 0 to 100 were presented as mean and 95% confidence interval (CI). In order to compare characteristics of the groups of participants according to the decisions met (Patient A, Patient B, or draw lot) we employed an analysis of variance (ANOVA) for comparing the mean age of participants and a binomial test for categorical variables such as gender, religion and health care profession.

## RESULTS

### Recall and demographic data

The social media campaign was launched on March 30, 2020. From April 1 to 12, 2020, the online survey was clicked a total of 3438 times. Overall, 1626 participants completed the entire survey (47%). Only complete surveys were included in this analysis.

Mean age of the participants was 40 years (standard deviation 14 years, median 39 years, range 12 to 80 years (table 1). Most participants were female (n=984, 61%), 623 were male and 4 considered themselves diverse, while 15 elected not to reveal their gender. A total of 645 (40%) worked in a medical or health profession.

**Table 1:** Demographics of the respondents compared to the German general population (Reference [10] except where noted). Respondends tended to be more females and slightly younger than the general population. Also, healthcare professionals were overrepresented in our collective.

| Collective (n) | Gender (n [%]) | Age (years) | Religion |  |  | Healthcare profession (%) |
| --- | --- | --- | --- | --- | --- | --- |
|  |  |  | Protestant (n, [%]) | Catholic (n, [%]) | Atheist (n, [%]) |  |
| Respondents study (1626) | Female 984 [61%]<br>Male 623 [38%]<br>Diverse 4 [<1%] | Mean 40<br>Median 39<br>Range 12 to 80 | 446 [29%] | 432 [28%] | 318 [21%] | 645 [40%] |
| General population (83.2m) | Female 42.1m [51%]<br>Male 41.0m [49%]<br>Diverse 150 [<1%]<br>[18] | Mean 44<br>Median 47<br>Range 0 - 113 | 21.1m [25%] | 23.0m [28%] | 34m - 40m [41-49%]<br>[19] | 5.7m [13%]<br>(of 44.8m employed) |

Abbreviations: m - million

The distribution of religious beliefs is also depicted in table 1. Most commonly participants were of protestant Christian faith (n=446 [29.2%]), and catholic Christian faith (n=432 [28.3%]), followed by atheists-agnostics (n=318 [20.8%]), and those that elected not to give any specifics regarding their religious denomination (n=247 [16.2%]). Less commonly represented were other Christian faith (n=67 [4.4%]), Islam (n=13 [0.9%]), Judaism (n=4 [0.3%]) and one Hindu person.

### Rationing decisions according to patient characteristics

Figure 1 illustrates the triage decisions. A majority of participants decided to provide the ventilator to the younger patient, the patient without co-morbidities, the patient with explicit consent, and the mother taking care of 4 children. The majority of participants elected to draw lots when *insurance status, asylum status/nationality*, and *social status* was in question. Nearly equal distribution for patient A, patient B and drawing lots was found for the criteria *chance of treatment success/prognosis*.

**Figure 1:**
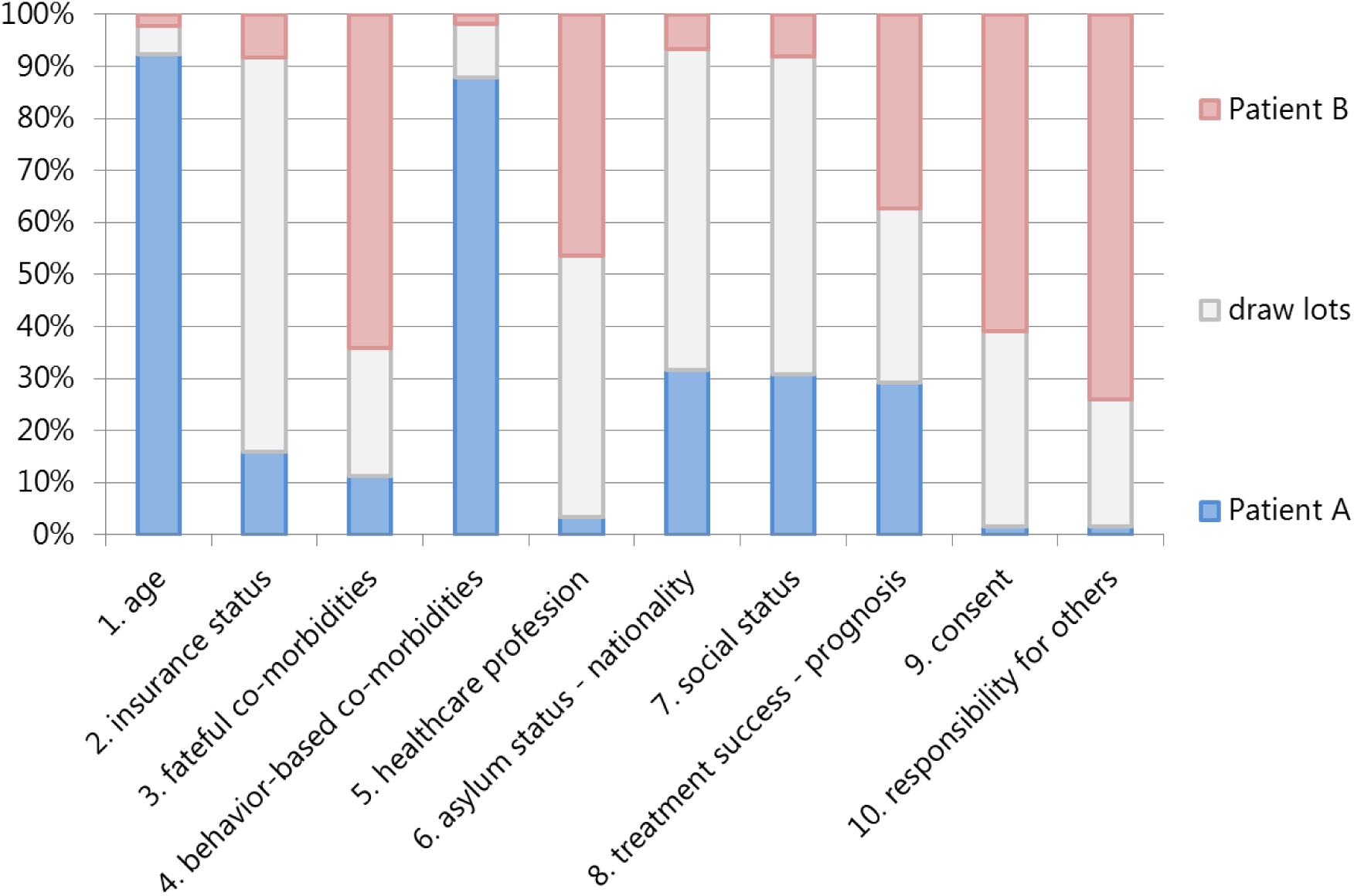
Participants’ decisions on the case scenarios. The paired characteristics of the imaginary patients A versus B comprise 1. young/old, 2. public/private insurance, 3. with or without fateful co-morbidities, 4. with or without behavior-associated co-morbidities, 5. working or not working/working in healthcare, 6. local nationality/asylum seeker, 7. high/low social status, 8. unfavorable/favorable prognosis, 9. consent given/lacking, 10. single household/mother of 4 children.

### Opinions on triage criteria

The relative weighting of the respective concrete triage criteria are found in figure 2. The participants scored *chance of treatment success/prognosis* as the most important criterion for rationing life-saving resources (mean score 77.8 [95%CI 76.7-78.8]). According to the participants’ opinions, *behavior-associated co-morbidities* (62.1 [95%CI 60.7-63.5]) should be as much of a criterion against receiving a scarce lifesaving therapy as *responsibility for others* should be a criterion in favor of receiving the therapy (62.1 [95%CI 60.8-63.6]). Also, participants generally favored younger over older patients (59.8 [95%CI 58.7-60.9]). On the other hand, according to the participants, *insurance status* (5.9 [95%CI 5.2-6.7]), *asylum status/nationality* (19.1 [95%CI 17.8-20.4]), as well as *social status* (24.8 [95%CI 23.4-26.2]) should not play a major role. *Fateful co-morbidities* (49.1 [95%CI 47.7-50.6]), *healthcare profession* (43.6 [95%CI 42.1-45.1]) and explicit *consent for intensive care* (52.6 [95%CI 51.1-54.1]) scored somewhat in the middle.

**Figure 2:**
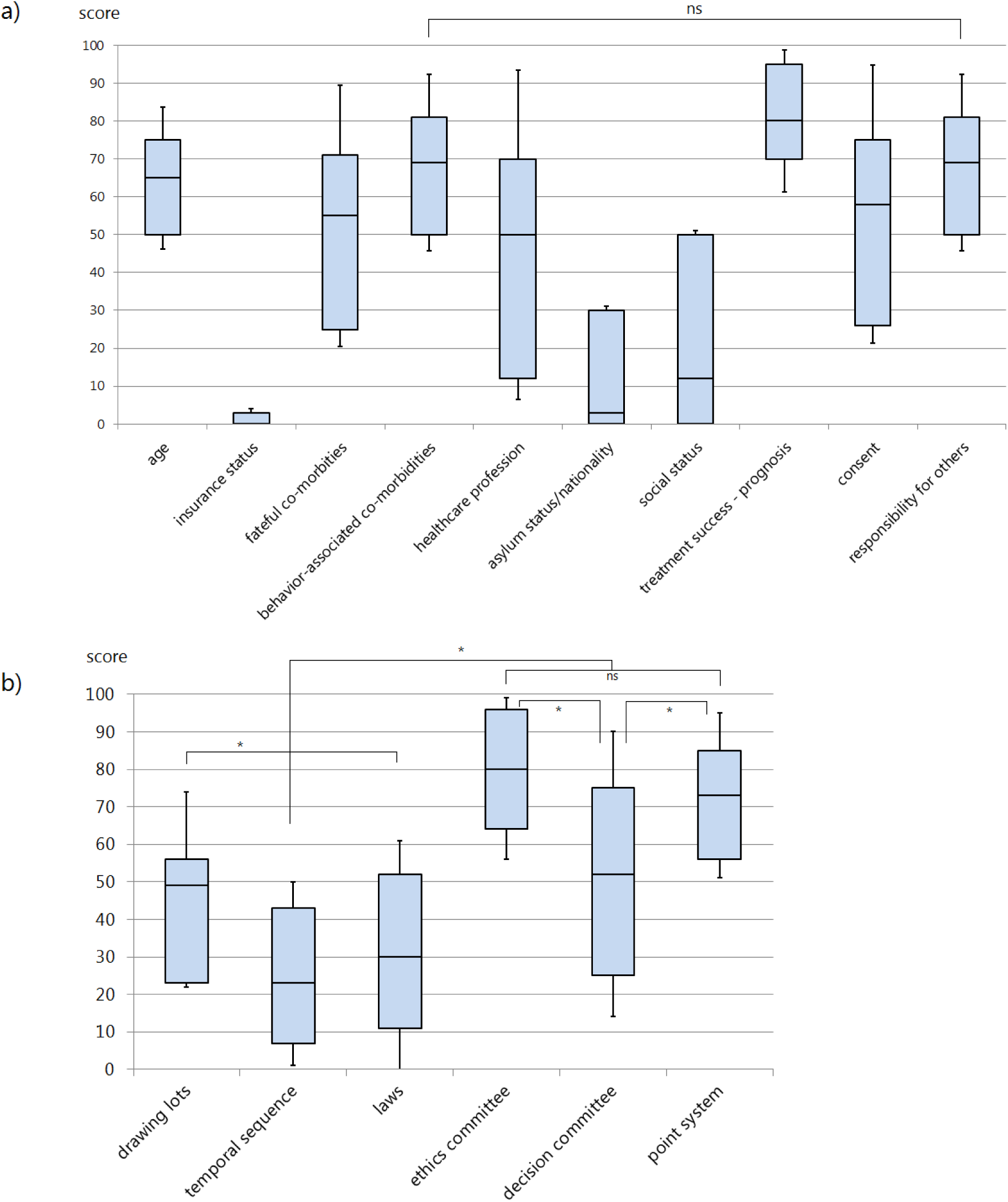
Opinions of the participants regarding the weighting of the respective triage criteria (a) and triage modalities (b). Participants regarded triage by prognosis as the most appropriate criterion, followed by behavior-associated co-morbidities and responsibility for others, as well as age (a). Most participants determined ethics committees and a point-system similar to that used in organ transplantation to be the most appropriate triage modalities (b). (ns: non-significant, *: p<0·05).

### Opinions on decision modalities

The participants viewed *ethics committees* that provide on-site interdisciplinary guidance to practicing physicians as the most helpful modality to make triage decisions (75.5 [95%CI 74.3-76.7]), followed by *point systems* similar to what is established for the distribution process in organ transplantation (68.5 [95%CI 67.3-69.7]). Triaging according to *temporal sequence* of presentation (27.3 [95%CI 26.1-28.5]) and *laws* (34.4 [95%CI 33.1-35.7]) were viewed as less helpful (figure 2). The scores regarding *drawing lots* (41.4 [95%CI 40.2-42.6]) and *decision committees* (50.5 [95%CI 49.0-52.0) were scored in the neutral middle.

### Demographics

Table 2 shows the associations of demographic factors with the responses. Older participants preferred drawing lots when the triage was based on *age*. Women preferred drawing lots for *fateful co-morbidities, healthcare profession*, as well as for *asylum status/nationality* and *social status*. Atheists-agnostics less often chose treating the patient with an *unfavorable prognosis* than participants of religious faiths. Working in healthcare almost always influenced the decision, except for the criteria *social status* and *healthcare profession*.

**Table 2:**
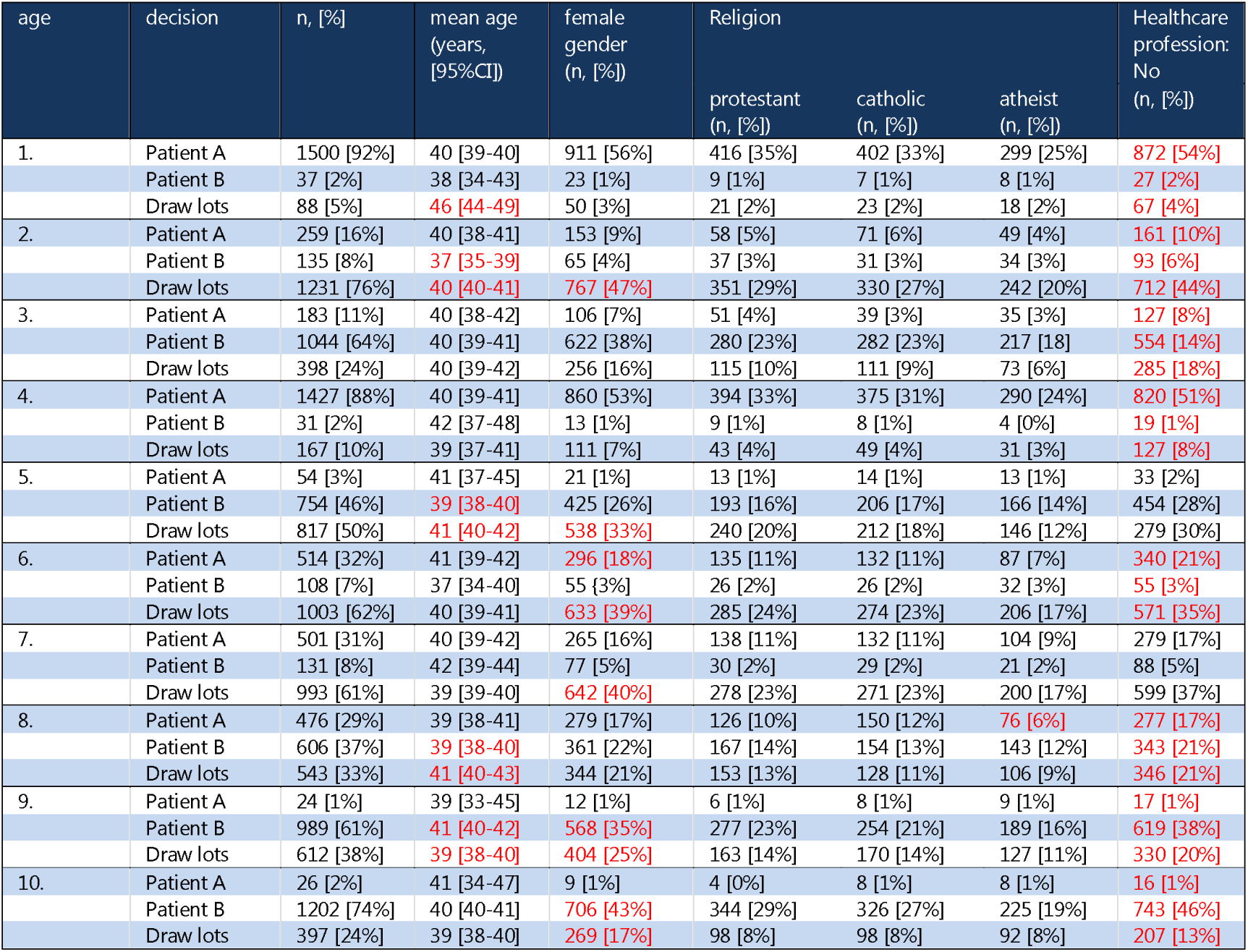
Overview of the responses according to age, gender, religious denomination, and working in a healthcare profession. Discrepancies to 100 are due to rounding. Significant differences are marked in red (regarding healthcare profession, red marks difference to those not working versus working in healthcare).

| age | decision | n, [%] | mean age<br>(years,<br>[95%CI]) | female<br>gender<br>(n, [%]) | Religion |  |  | Healthcare<br>profession:<br>No<br>(n, [%]) |
| --- | --- | --- | --- | --- | --- | --- | --- | --- |
|  |  |  |  |  | protestant<br>(n, [%]) | catholic<br>(n, [%]) | atheist<br>(n, [%]) |  |
| 1. | Patient A | 1500 [92%] | 40 [39-40] | 911 [56%] | 416 [35%] | 402 [33%] | 299 [25%] | 872 [54%] |
|  | Patient B | 37 [2%] | 38 [34-43] | 23 [1%] | 9 [1%] | 7 [1%] | 8 [1%] | 27 [2%] |
|  | Draw lots | 88 [5%] | 46 [44-49] | 50 [3%] | 21 [2%] | 23 [2%] | 18 [2%] | 67 [4%] |
| 2. | Patient A | 259 [16%] | 40 [38-41] | 153 [9%] | 58 [5%] | 71 [6%] | 49 [4%] | 161 [10%] |
|  | Patient B | 135 [8%] | 37 [35-39] | 65 [4%] | 37 [3%] | 31 [3%] | 34 [3%] | 93 [6%] |
|  | Draw lots | 1231 [76%] | 40 [40-41] | 767 [47%] | 351 [29%] | 330 [27%] | 242 [20%] | 712 [44%] |
| 3. | Patient A | 183 [11%] | 40 [38-42] | 106 [7%] | 51 [4%] | 39 [3%] | 35 [3%] | 127 [8%] |
|  | Patient B | 1044 [64%] | 40 [39-41] | 622 [38%] | 280 [23%] | 282 [23%] | 217 [18%] | 554 [34%] |
|  | Draw lots | 398 [24%] | 40 [39-42] | 256 [16%] | 115 [10%] | 111 [9%] | 73 [6%] | 285 [18%] |
| 4. | Patient A | 1427 [88%] | 40 [39-41] | 860 [53%] | 394 [33%] | 375 [31%] | 290 [24%] | 820 [51%] |
|  | Patient B | 31 [2%] | 42 [37-48] | 13 [1%] | 9 [1%] | 8 [1%] | 4 [0%] | 19 [1%] |
|  | Draw lots | 167 [10%] | 39 [37-41] | 111 [7%] | 43 [4%] | 49 [4%] | 31 [3%] | 127 [8%] |
| 5. | Patient A | 54 [3%] | 41 [37-45] | 21 [1%] | 13 [1%] | 14 [1%] | 13 [1%] | 33 [2%] |
|  | Patient B | 754 [46%] | 39 [38-40] | 425 [26%] | 193 [16%] | 206 [17%] | 166 [14%] | 454 [28%] |
|  | Draw lots | 817 [50%] | 41 [40-42] | 538 [33%] | 240 [20%] | 212 [18%] | 146 [12%] | 279 [30%] |
| 6. | Patient A | 514 [32%] | 41 [39-42] | 296 [18%] | 135 [11%] | 132 [11%] | 87 [7%] | 340 [21%] |
|  | Patient B | 108 [7%] | 37 [34-40] | 55 [3%] | 26 [2%] | 26 [2%] | 32 [3%] | 55 [3%] |
|  | Draw lots | 1003 [62%] | 40 [39-41] | 633 [39%] | 285 [24%] | 274 [23%] | 206 [17%] | 571 [35%] |
| 7. | Patient A | 501 [31%] | 40 [39-42] | 265 [16%] | 138 [11%] | 132 [11%] | 104 [9%] | 279 [17%] |
|  | Patient B | 131 [8%] | 42 [39-44] | 77 [5%] | 30 [2%] | 29 [2%] | 21 [2%] | 88 [5%] |
|  | Draw lots | 993 [61%] | 39 [39-40] | 642 [40%] | 278 [23%] | 271 [23%] | 200 [17%] | 599 [37%] |
| 8. | Patient A | 476 [29%] | 39 [38-41] | 279 [17%] | 126 [10%] | 150 [12%] | 76 [6%] | 277 [17%] |
|  | Patient B | 606 [37%] | 39 [38-40] | 361 [22%] | 167 [14%] | 154 [13%] | 143 [12%] | 343 [21%] |
|  | Draw lots | 543 [33%] | 41 [40-43] | 344 [21%] | 153 [13%] | 128 [11%] | 106 [9%] | 346 [21%] |
| 9. | Patient A | 24 [1%] | 39 [33-45] | 12 [1%] | 6 [1%] | 8 [1%] | 9 [1%] | 17 [1%] |
|  | Patient B | 989 [61%] | 41 [40-42] | 568 [35%] | 277 [23%] | 254 [21%] | 189 [16%] | 619 [38%] |
|  | Draw lots | 612 [38%] | 39 [38-40] | 404 [25%] | 163 [14%] | 170 [14%] | 127 [11%] | 330 [20%] |
| 10. | Patient A | 26 [2%] | 41 [34-47] | 9 [1%] | 4 [0%] | 8 [1%] | 8 [1%] | 16 [1%] |
|  | Patient B | 1202 [74%] | 40 [40-41] | 706 [43%] | 344 [29%] | 326 [27%] | 225 [19%] | 743 [46%] |
|  | Draw lots | 397 [24%] | 39 [38-40] | 269 [17%] | 98 [8%] | 98 [8%] | 92 [8%] | 207 [13%] |

### Decision certainty

All participants together were more insecure about their decisions regarding age and *behavior-associated co-morbidities* in comparison to other factors (table 3). Female participants were generally more secure about their decisions than males. Atheists/agnostics were generally less secure about their decisions than protestant or catholic Christians.

**Table 3:** Feeling of security about the decision according to participant characteristics (scale from 0 to 100). Statistical significant values marked in red.

| case | mean | 95% CI |
| --- | --- | --- |
| 1. age | 38 | 36-39 |
| 2. insurance status | 46 | 44-48 |
| 3. fateful co-morbidities | 45 | 44-46 |
| 4. behavior-associated co-morbidities | 37 | 35-38 |
| 5. healthcare profession | 48 | 46-49 |
| 6. asylum status - nationality | 45 | 43-46 |
| 7. social status | 45 | 44-47 |
| 8. chance of treatment success - prognosis | 42 | 41-44 |
| 9. consent | 46 | 45-48 |
| 10. responsibility for others | 41 | 40-43 |
| male | 41 | 40-43 |
| female | 45 | 44-46 |
| diverse | 54 | 12-97 |
| healthcare profession: yes | 44 | 42-45 |
| healthcare profession: no | 43 | 42-44 |
| protestant Christian | 45 | 43-47 |
| catholic Christian | 45 | 44-47 |
| atheist-agnostic | 40 | 38-42 |
| other | 41 | 39-43 |

## DISCUSSION

To our knowledge, this is the first empirical survey study on triage attitudes of a cross-sectional population sample using social media distribution. This study shows that using this methodology, a relatively large sample of participants can be recruited in a short amount of time, thus promoting participatory processes so essential in democratic societies, particularly in the context of an ongoing crisis. This study gives insight into opinions and attitudes of a select sample of the German population on rationing life-saving therapies during the European peak of the corona-pandemic. It seems noteworthy, that during this peek, the rationing of scarce resources and triage was highly present in German (mass-) media, particularly with reference to the devastating situation in Northern Italy, parts of France, Spain, and most currently, hot-spots in the United States, where physicians were suddenly put in charge of what figuratively could be described as *a Titanic with not enough life-boats*. This led to a situation in which most Germans were at least confronted with – if not interested in – the concept of triage which they may have never heard of before. This was also one of the reasons for addressing the issue at the first European peak of the pandemic and sending out the questionnaire in a timely manner, using the window of opportunity to generate knowledge about values and beliefs in the general population to avoid the implementation of morally counterintuitive or socially inacceptable procedures in the process of preparing for the impact of the COVID-19 pandemic.

The word triage etymologically stems from the French verb trier, which means to sort or select. The principle of categorizing injured soldiers on the battleground according to their treatability was developed by the Napoleonic military physician Baron Dominique-Jean Larrey (1766-1842).[11] Larrey also invented the so-called *ambulances volantes*, with which injured soldiers could be transported rapidly to the field lazaret. Once there, they were triaged according to acuity and prognosis, and received tailored treatment. The COVID-19 crisis has brought the concept of triaging scarce healthcare resources into the public discourse.[12]

Triage decisions are usually made according to utilitarian principles, so according to how the scarce resources must be distributed to afford most beneficence for the greatest number of patients. Superficially, this translates into the goal of simply saving the highest number of lives. However, upon closer examination, it becomes more complex. Should a young person be treated the same as a 93 year old? From a utilitarian perspective, one could argue that by favoring the younger patient, more life-years are saved. Should a patient in a vegetative state without a prospect of improvement receive the same chance of survival as a healthy individual who is mentally normal? With other words, in a difficult triage situation, should really every human life be valued and treated equally? The German ethics council demands that egalitarian principles should be followed, based on the dignity of every human person.[13] What appears obvious on first glance becomes challenging and counterintuitive in reality. Consequently, one can hardly blame physicians working in SARS-CoV-2 hotspots for including expected life-years and quality of life in their ad-hoc decisions of distributing lifesaving resources. Some authorities actually explicitly demand taking these factors into account.[14] From a socioeconomic standpoint, it may also be desirable for a society to prefer younger, more active and fertile individuals in the triage of lifesaving measures. In this sense, the call for “women and children first” may as well be regarded as a - possibly cynical - neoliberal attempt of pushing an otherwise unacceptable construct of human economic value.

On engaging deeper with this subject, one inevitably encounters John Rawls’ contractual approach,[15] in which the members of a society agree upon fair, universal rules behind an imaginary veil of ignorance. In our case, these rules pertain to the criteria which should be applied for triage in the event of a disaster such as the COVID-19 pandemic. The model is based on the premise that nobody knows which position one would occupy in such a situation. One could be anybody, the infant or the elderly individual, the rich or the poor, the smoker or the health fanatic. This study tries to emulate such a paradigm by presenting the different scenarios and allowing the participant to decide from an outside, noninvolved standpoint.

Nonetheless, it needs to be stressed that triage criteria should not be considered universal in the way it is usually understood in ethics, but varying according to the moral principles and priorities of a given society. While they must be adapted to a certain situation, this should not lead to complete moral relativism that is a fully fledged particularism, which may also be found as an argumentative position in ethical debates, particularly in very diverse societies. To facilitate a more pragmatic decision-making in the realm of clinical ethics, the results of this study must be viewed in light of basic, universal, and associative norms embedded in a larger framework of ethical principles, human dignity, and human rights in the sense of integrative particularisms.

Another aspect is how the triage should be conducted, what modality should be used. In the acute phase of a disaster, patients are often treated by chronologic sequence. Patients who arrive to the hospital earlier are treated first. As long as resources are available for all, this approach is fair. Once the available capacity is surpassed, however, this distribution becomes arbitrary and inherently unfair. Drawing lots provides every patient with the same chance of receiving therapy, however, this approach is only legitimate in patients presenting under the same circumstances and with the same prognosis, a prerequisite that is hardly imaginable in clinical practice. Acting upon set laws predetermined by the legislative alone would provide legal security, but strongly encroach on individual and situational treatment decisions. Expert panels that decide upon treatment allocation remotely have been criticized polemically as “death panels” in the context of the implementation of the Affordable Care Act in the United States.[16] In most western countries, ethics committees in the form of interdisciplinary advisory teams are available to consult on and solve complex triage dilemmas, while preserving autonomy of the patient and treating physicians.

The participants of this study most vehemently based their triage decisions on prognosis, age and responsibility for others. They also regarded behavior-associated co-morbidities as a negative criterion for receiving therapy. On the other hand, an overwhelming majority rejected insurance, social, and asylum status as factors in the triage process.

Concerning the triage modality, most were in favor of ethics committees and a point system. These are already commonplace in modern healthcare culture, as well as organ transplant systems. Since healthcare professionals were relatively overrepresented in the study sample, the responses can be interpreted as an affirmation of the existing triage mechanisms.

An additional interesting finding of our study is the influence of demographics. Older participants did not universally favor the older imaginary patients, but were more likely to draw lots. Women tended to employ randomization in questions of status more frequently, and overall seemed surer about their decision than men. Christians also expressed more certainty about their decisions than atheists/agnostics. This may be the result of an internalized religious moral framework that could help making triage decisions. The sample size of the other religions was too small to draw conclusions.

Since the beginning of the COVID-19 spread, several associations have published guidelines on triage. One of these was the German Interdisciplinary Association for Intensive Care and Emergency Medicine (Deutschen Interdisziplinären Vereinigung für Intensiv-und Notfallmedizin, DIVI).[17] In these guidelines, the authors focus on the role of prognosis and consent of the patient and postulate that triage decisions should be made in consensus by the individuals involved in care. Aborting treatment of one patient in the interest of another is not admissible because of the German constitution, which highlights the dignity of every individual as equal. Also, prioritizing solely by age is also unacceptable. Other concrete criteria are not mentioned in this publication.

Our study provides an initial impulse to substantiate triage criteria in the context of the COVID-19 pandemic. By using case-based scenarios, we tried to include an emotional component in the decision process, beyond the participants’ abstract views on triage criteria.

However, it also has several limitations. The most obvious one is the fact that an online survey based on recruitment through social media does not necessarily confer a representative sample of society in general. In addition, those working in healthcare seemed overrepresented. We cannot make any statements regarding recall, since the denominator of how many people actually were reached is lacking. What we have shown, however, is that a large group of people can be recruited through an online survey in a very short time, and that the interest was high enough so that almost half of those that clicked the link also completed the survey in whole. We interpret this finding as a general interest in the subject of triage, and a willingness to participate in the public discourse around the factors that should play a role. In the future, more precise and representative measurements may be obtained by using more elaborate distribution channels, focus groups, and more detailed demographic information.

Also, one may question whether the general public should be included in the discussion about triage criteria or not. While some may argue that categorical principles interpreted by experts in the relevant fields should receive priority, we strongly believe that it is appropriate and necessary in a democratic society to develop such criteria in a societal discourse and to facilitate participatory processes. In this regard, this study is also a small pilot test case for the implementation of such participation. Since a large part of this public exchange currently takes place on social media, we found it reasonable to evaluate whether these new technologies can be harnessed to foster a culture of participation and inclusion, even when the subject matter are complex ethical decisions. This study also shows that something as abstract as moral attitudes involved in medical triage can be quantified in an empirical study.

We suggest that the findings of this study are used as a backdrop for a broader public debriefing on triage criteria after the COVID-19 pandemic subsides. This should include a push to establish morally acceptable triage criteria and algorithms for future disasters and pandemics.

## CONCLUSION

This study generated a snapshot of the attitudes of a sample of German-speaking participants regarding criteria and modalities for the allocation of life-saving, limited resources during the height of the COVID-19 pandemic. According to the participants, prognosis, responsibility towards others, the absence of behavior-associated co-morbidities, along with younger age should be criteria for allocation of scarce ventilators. Insurance status, social status, as well as nationality and asylum status should not play a relevant role. Ethics-committees and a point system similar to what is used for organ transplantation were deemed potentially helpful in triage decision making. The findings and results of this study may be useful in creating future surveys addressed at a broader public. Similar but refined methods may be useful to expand future triage-algorithms by a dimension of societal consensus.

## Data Availability

All data relevant to the study is available upon reasonable request from the corresponding author.

## Contributors

Oliver J. Muensterer: Conception of the study; study design; creation of the online survey; data collection and processing; creating figures and tables; drafting and editing of the manuscript.

Emilio A. Gianicolo: Data processing; statistical analysis; editing tables; review and editing of manuscript.

Norbert W. Paul: Mentoring and overseeing the master thesis; conception of the study; study design; co-drafting, review and editing of the manuscript.

## Patient consent for publication

Not required. According to state law, institutional review board approval was waived because no patient data was included in the study. Participation in the online study was voluntary and anonymous. No identifiable information was collected or stored.

## Acknowledgements

Thanks to Grigorios Pilidis and Diana Walz for helping with the online survey on Lime. Thanks to the many participants in the survey that made this study possible.

## Data sharing Statement

All data relevant to the study is available upon reasonable request from the corresponding author.

## Appendix A: Online-survey

(translated from German, the original survey in German available upon request)

### Survey

Entry requested by participant marked in yellow

### Introduction

Imagine you are working as a physician in an intensive care unit of a hospital. In the following imaginary scenarios, a situation is described in which you will have to take a difficult decision.

You have two very ill COVID-19 patients who only have a chance of survival if they receive mechanical ventilation (a respirator). You only have one ventilator machine available. You must therefore make a decision on who receives the ventilator. You can also draw lots, allowing chance to decide who receives the ventilator. In that case, both patients have equal chances.

How would you decide? How sure are you about your decision? How appropriate do you think the particular characteristic is to determine who receives the ventilator?

This survey includes 10 of these scenarios. At the end of the survey, you will be asked a few questions regarding your age, gender, religious beliefs, and whether you work in healthcare. In total, the survey will take around 20 to 30 minutes.

By clicking Continue below you consent to participating in this study.

Continue

### 1. Case number 1

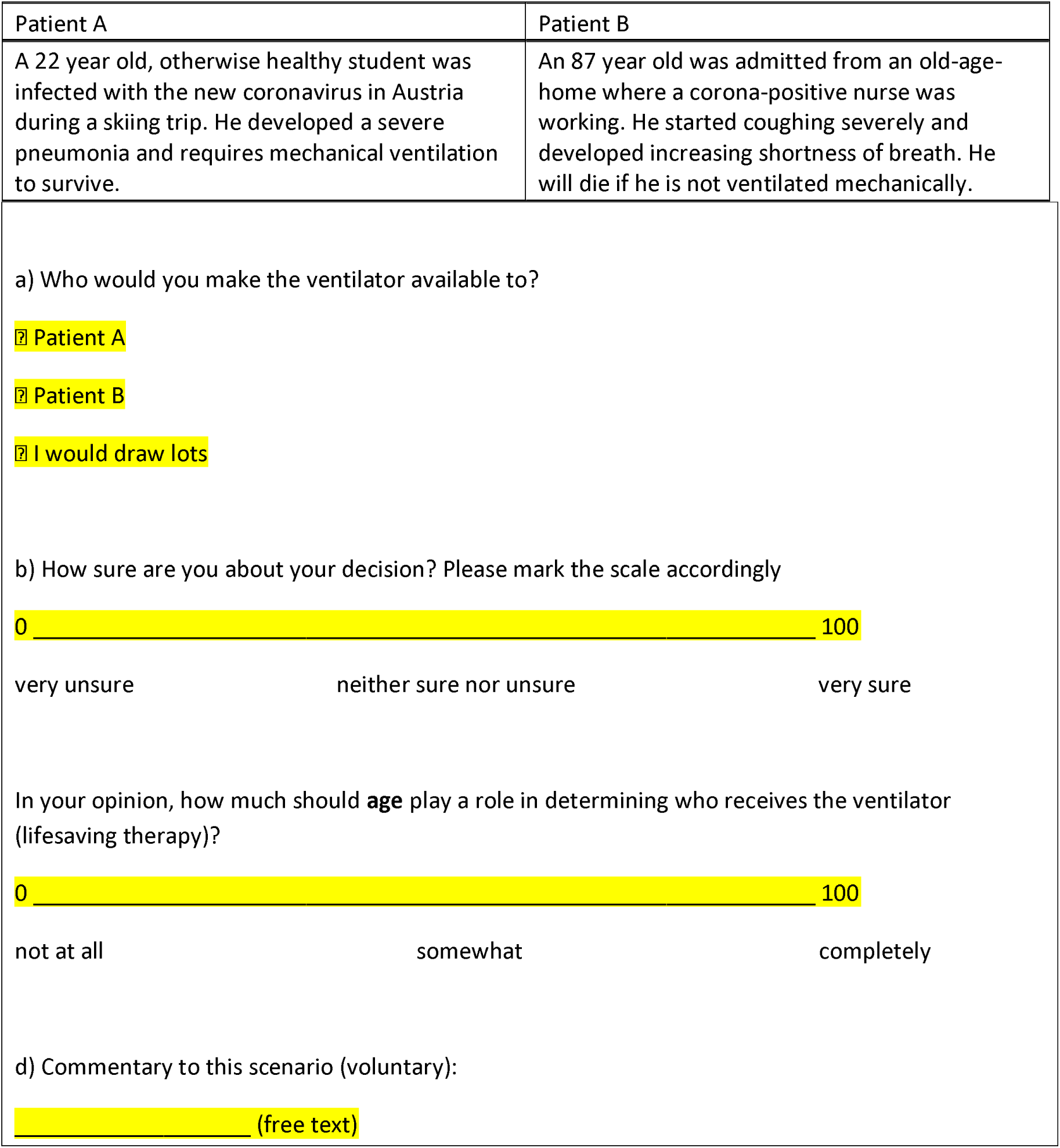

### 2. Case number 2

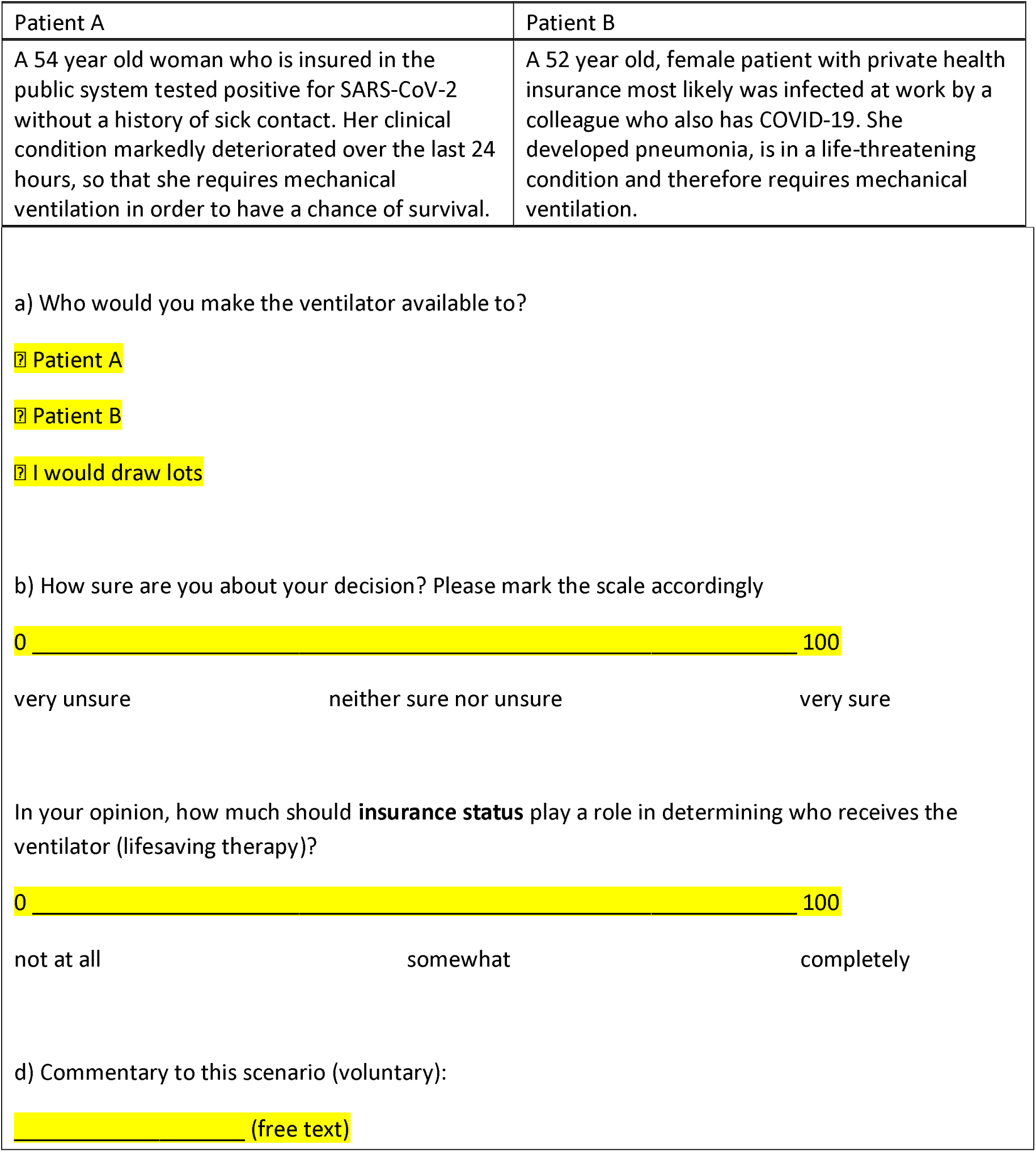

### 3. Case number 3

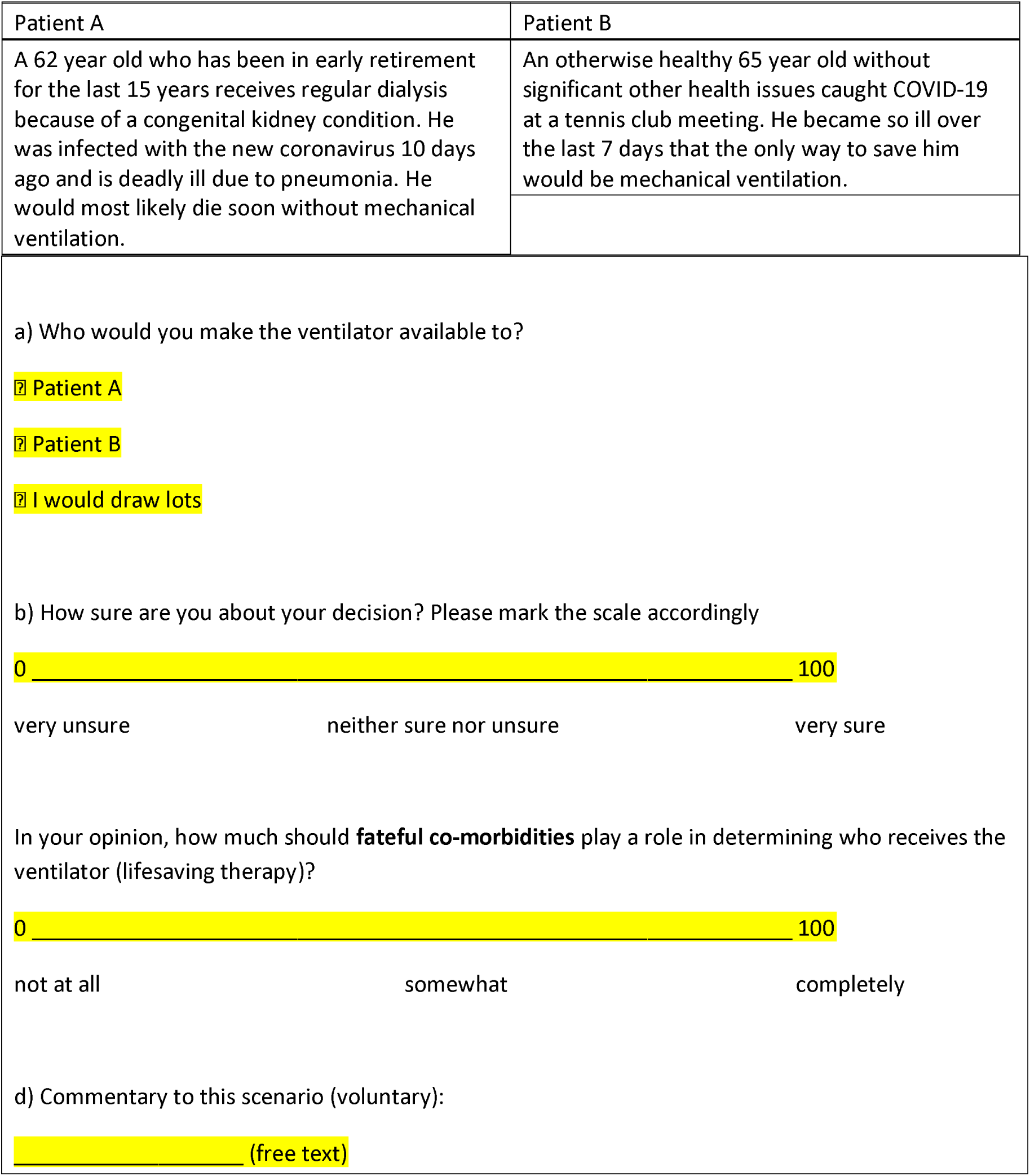

### 4. Case number 4

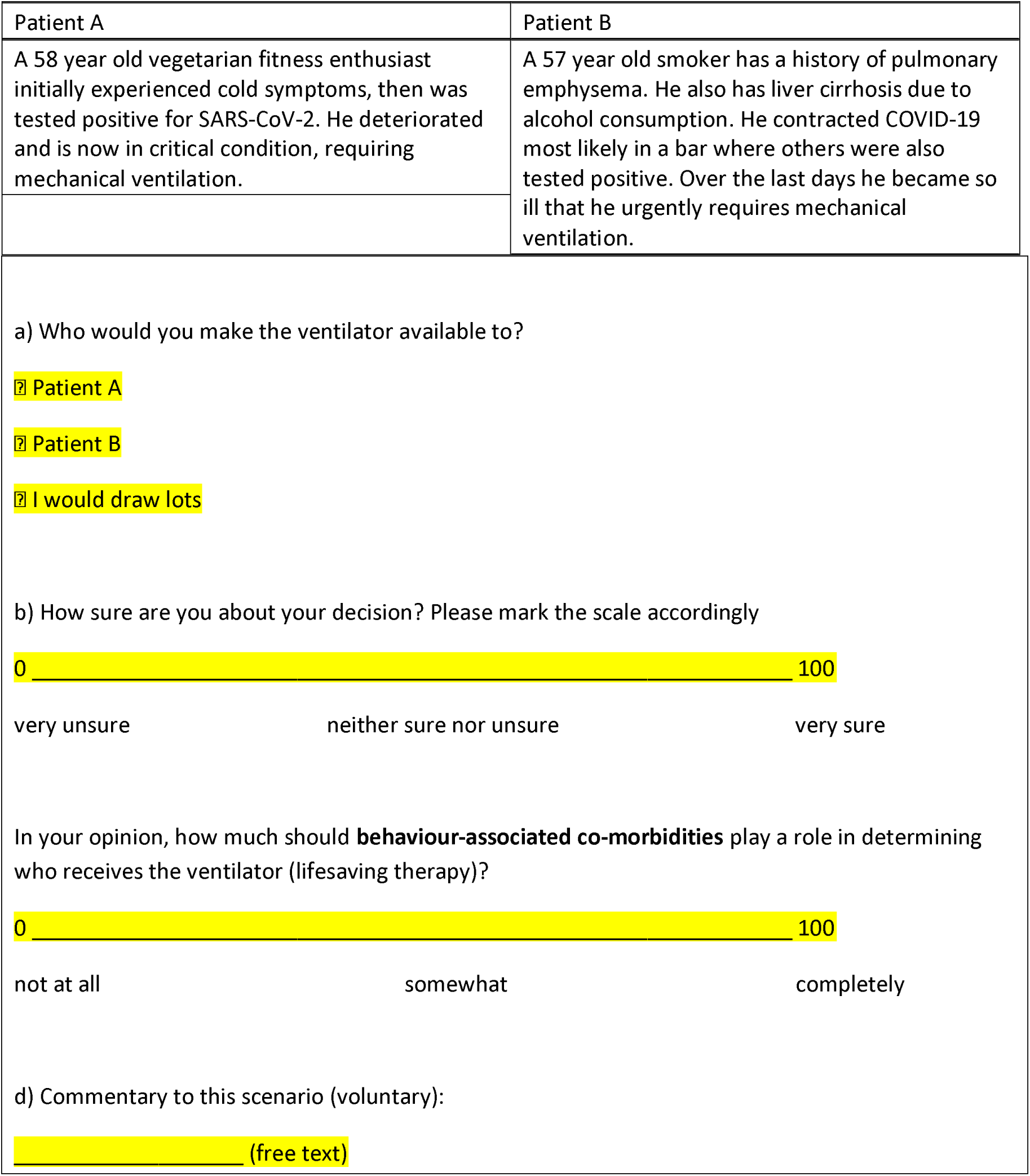

### 5. Case number 5

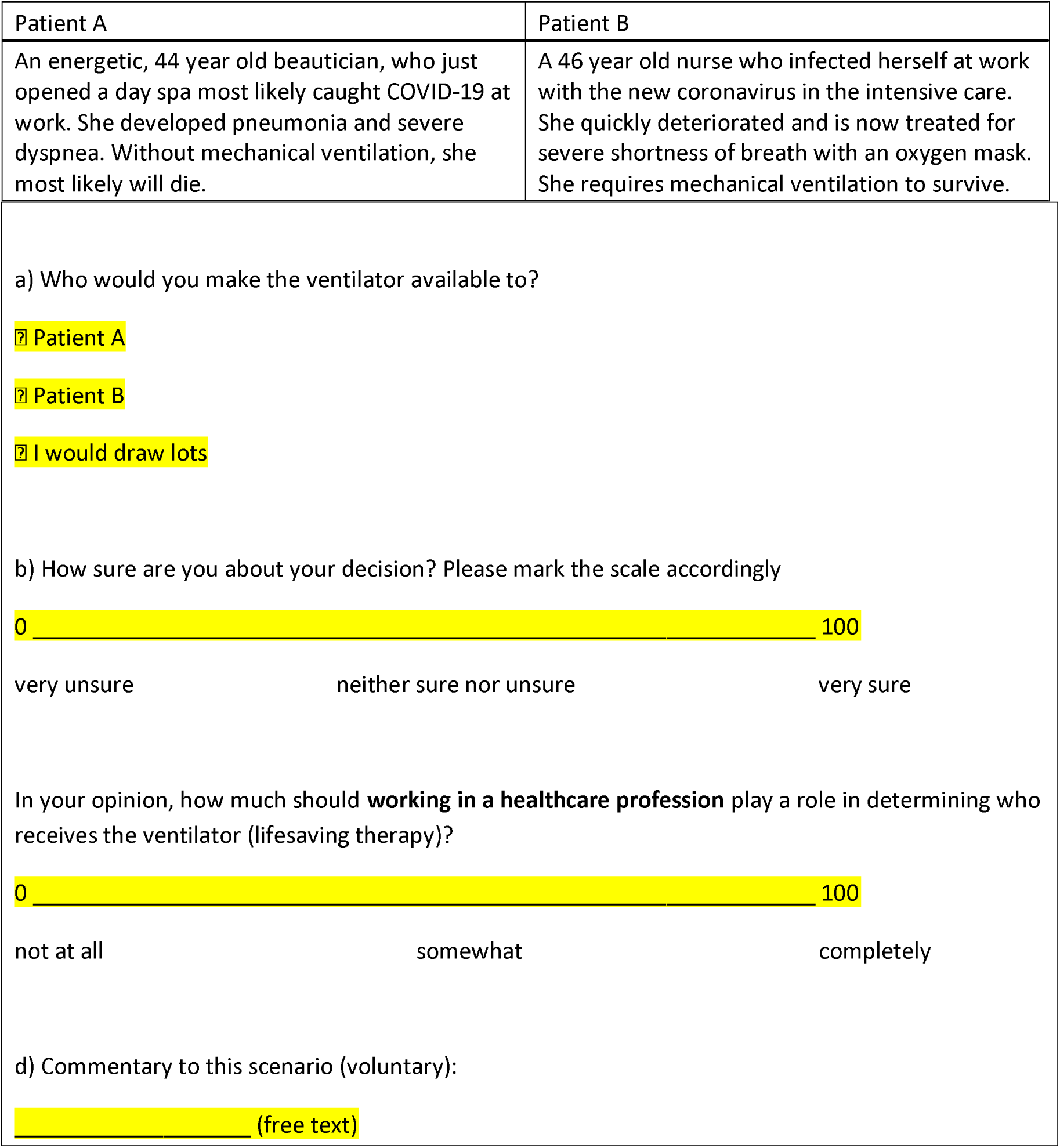

### 6. Case number 6

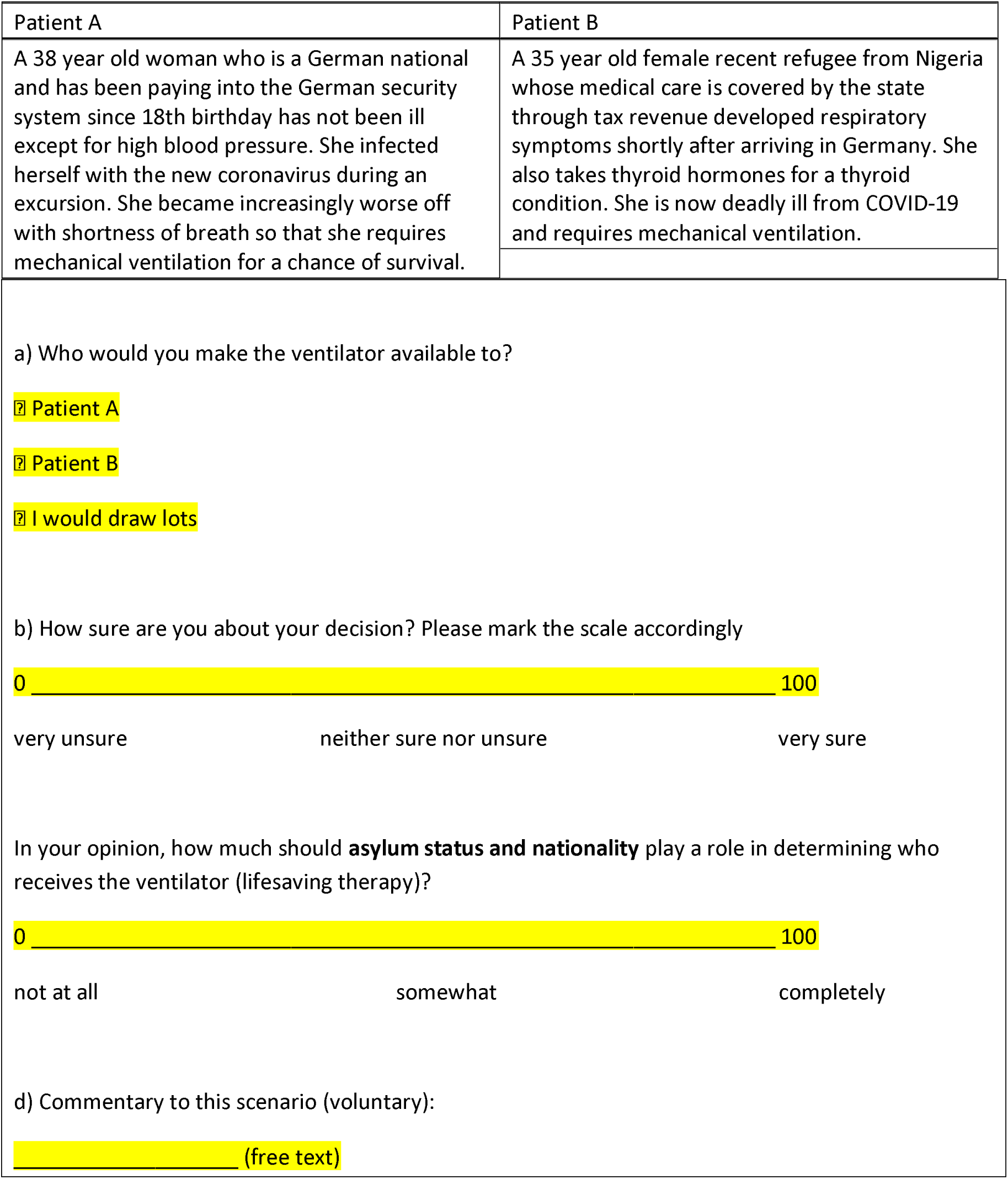

### 7. Case number 7

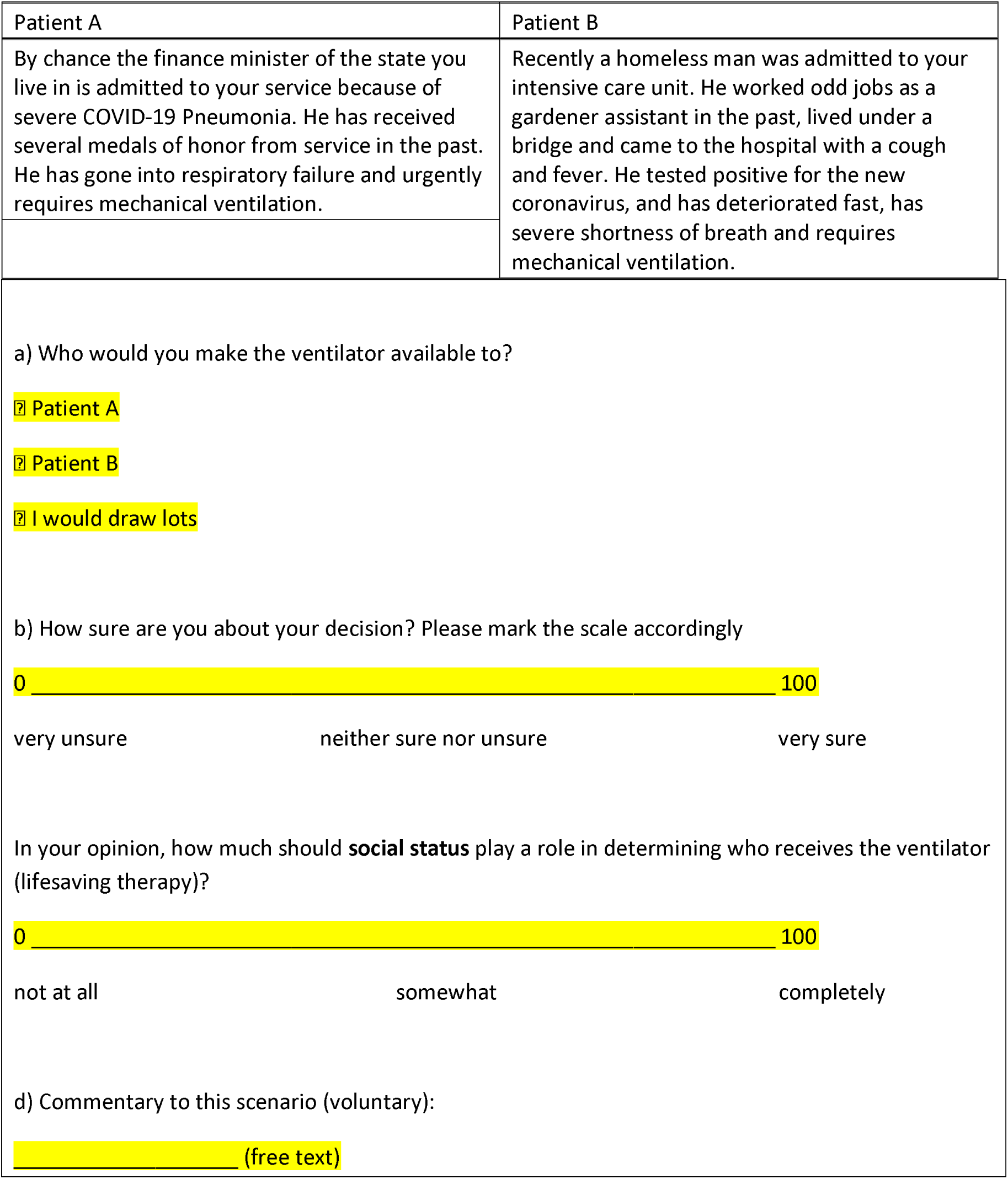

### 8. Case number 8

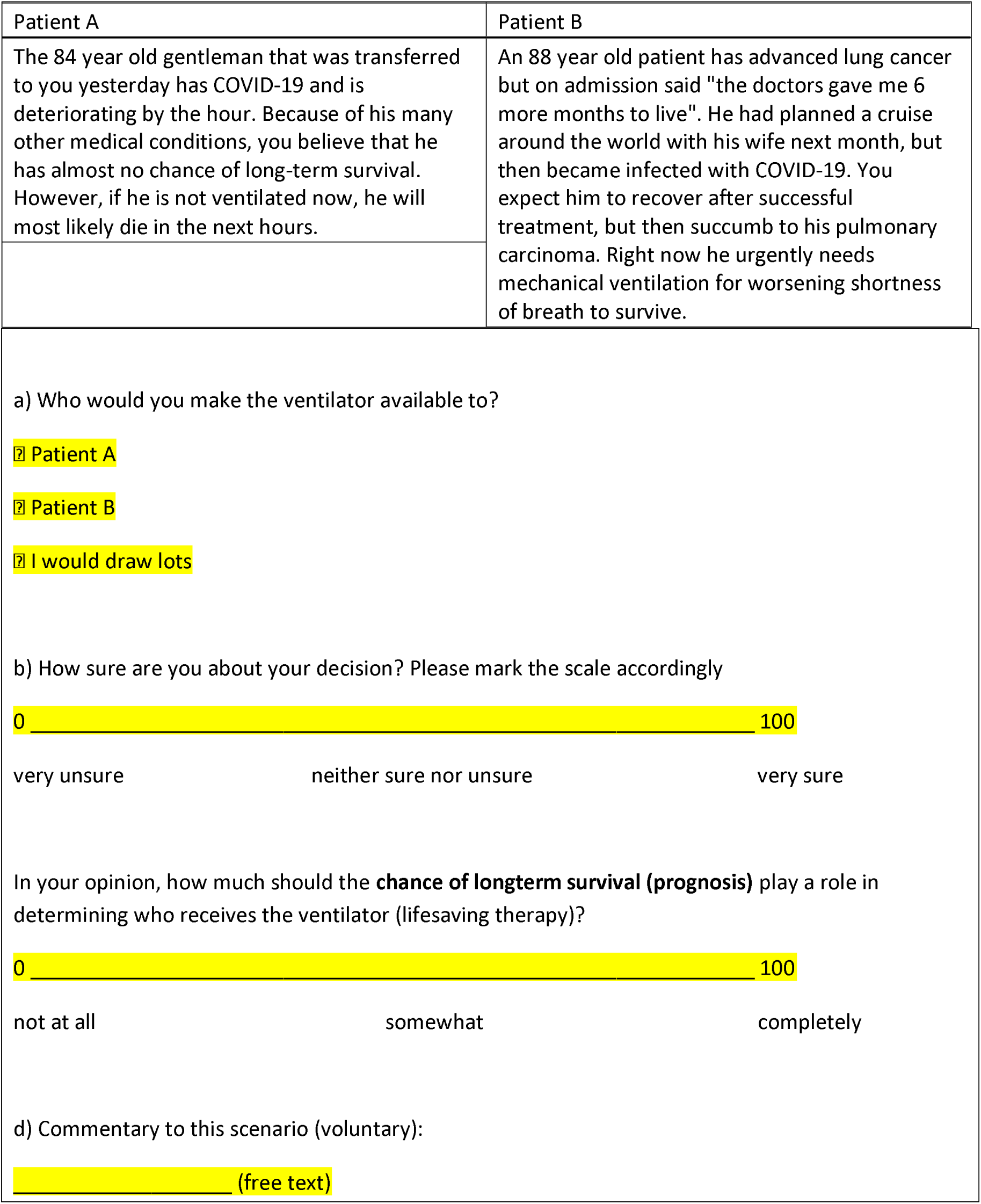

### 9. Case number 9

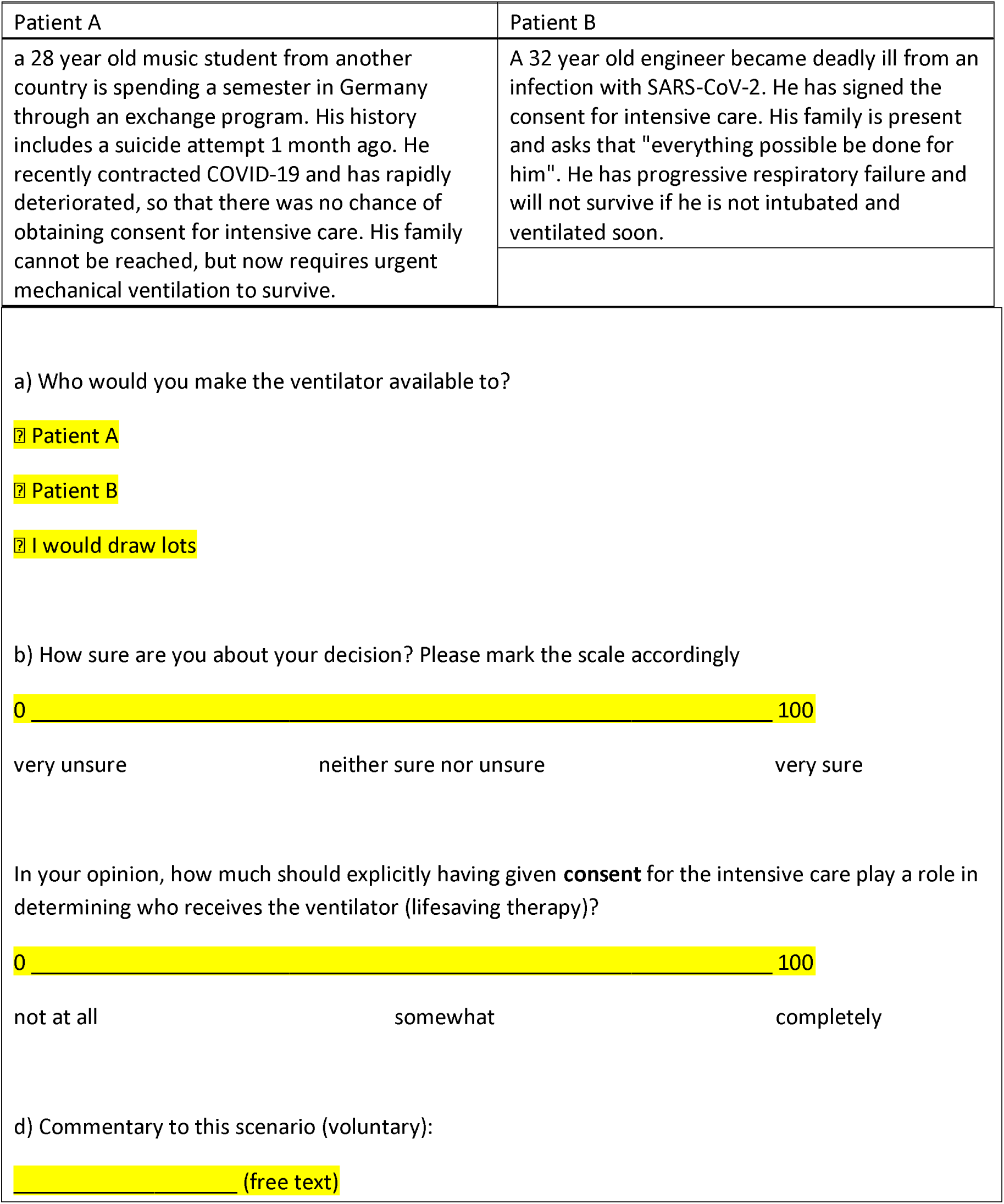

### 10. Case number 10

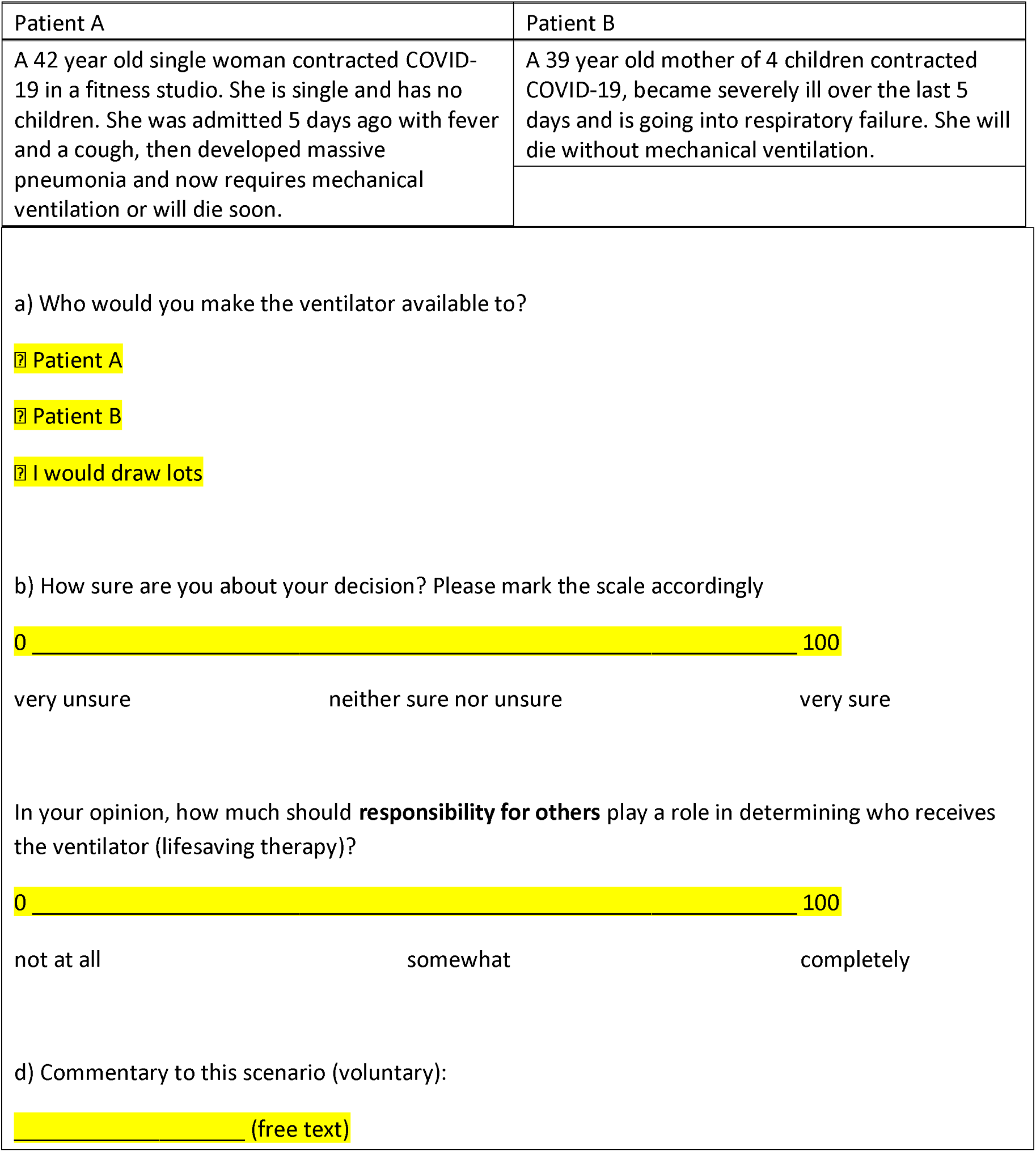
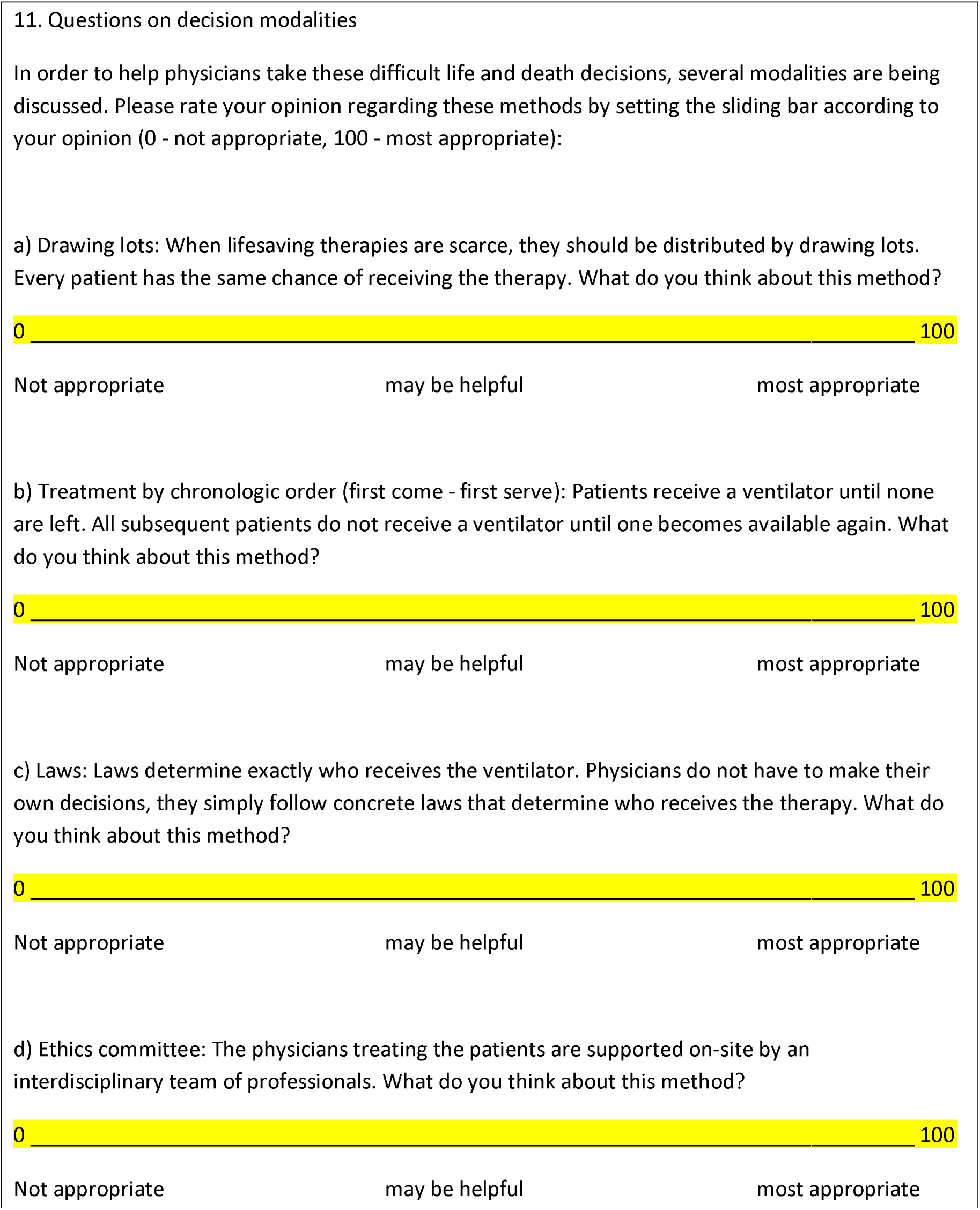
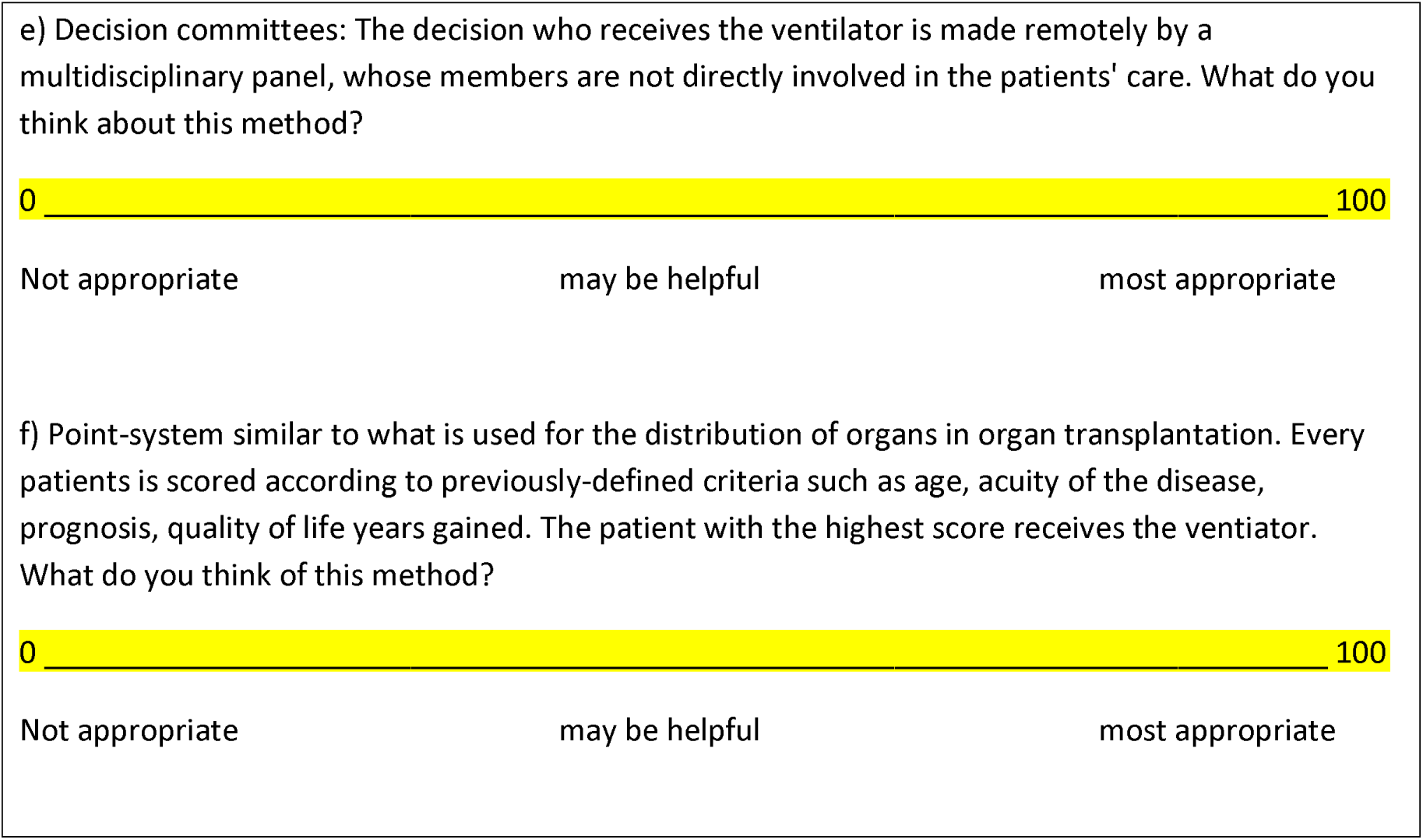
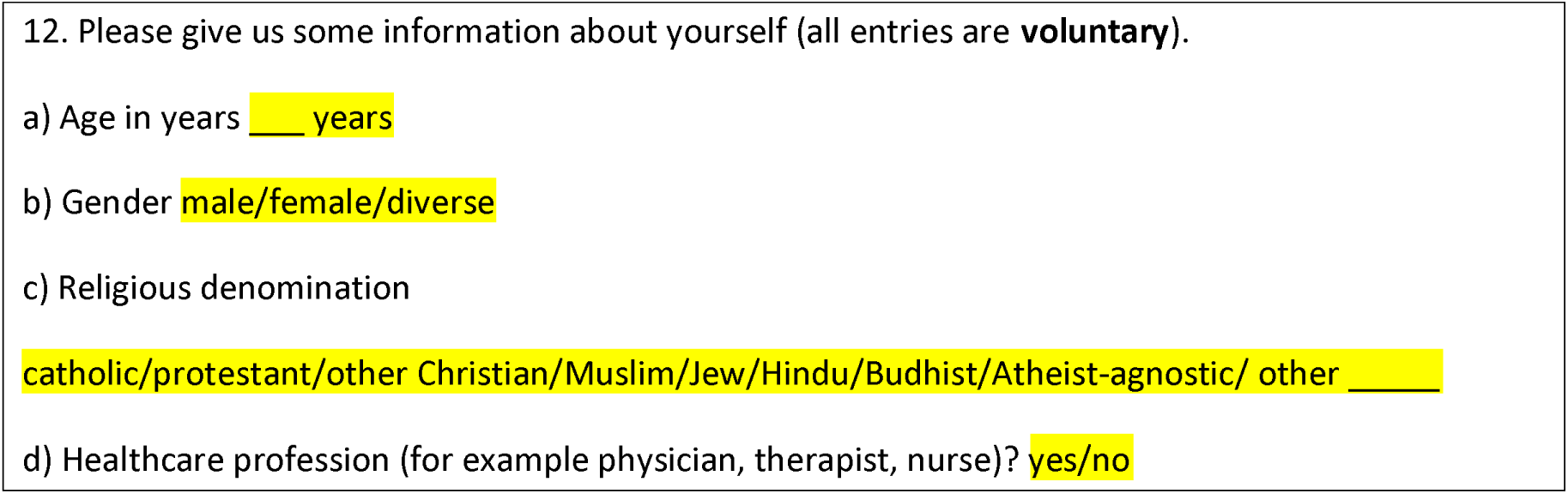

## REFERENCES

1 Rosenbaum L. Facing Covid-19 in Italy - Ethics, Logistics, and Therapeutics on the Epidemic’s Front Line. N Engl J Med 2020. doi:10.1056/NEJMp2005492. PMID: 32187459

2 Truog RD, Mitchell C, Daley GQ. The Toughest Triage - Allocating Ventilators in a Pandemic. N Engl J Med 2020. doi:10.1056/NEJMp2005689. PMID: 32202721

3 Remuzzi A, Remuzzi G: Covid-19 and Italy: what next? Lancet 2020:S0140-6736(20)30627-9

4 Nakao H, Ukai I, Kotani J. A review of the history of the origin of triage from a disaster medicine perspective. Acute Med Surg 2017;4:379–84

5 Simon R, Teperman S. The World Trade Center Attack: Lessons for disaster management. Crit Care 2001;5:318–20

6 Lübbe, Weyma: Corona Triage: A Commentary on the Triage Recommendations by Italian SIAARTI Medicals Regarding the Corona Crisis, VerfBlog, 2020/3/16, https://verfassungsblog.de/corona-triage-2/, DOI: https://doi.org/10.17176/20200317-002812-0 (accessed 27 July 2020)

7 Zimmermann T. Ärzte in Zeiten von Corona. Wer stirbt zuerst? Legal Tribune Online. https://www.lto.de/recht/hintergruende/h/corona-triage-tod-strafrecht-sterben-krankenhaus-entscheidung-auswahl/ (accessed 27 July 2020)

8 Hübner J, Schewe DM, Katalinic A, et al. Rechtsfragen der Ressourcenzuteilung in der COVID-19-Pandemie – Zwischen Utilitarismus und Lebenswertindifferenz [Legal Issues of Resource Allocation in the COVID-19 Pandemic - Between Utilitarianism and Life Value Indifference] Dtsch Med Wochenschr 2020;145:687–92

9 Mannelli C. Whose life to save? Scarce resources allocation in the COVID-19 outbreak. J Med Ethics 2020;46:364–6

10 Statistisches Bundesamt https://www.destatis.de/DE/Home/_inhalt.html (accessed 27 July 2020)

11 Brewer LA 3rd. Baron Dominique Jean Larrey (1766-1842). Father of modern military surgery, innovater, humanist. J Thorac Cardiovasc Surg 1986;92:1096–8

12 Schwartz K. Triage in Deutschland - Wer wird beatmet? Wer nicht? www.tagesschau.de/inland/corona-triage-intensivmedizin-101.html (accessed 21 July 2020)

13 German Ethics Council. Deutscher Ethikrat. Solidarity and Responsibility during the Coronavirus Crisis. https://www.ethikrat.org/fileadmin/Publikationen/Ad-hoc-Empfehlungen/englisch/recommendation-coronavirus-crisis.pdf (accessed 27 July 2020)

14 Persad G, Wertheimer A, Emanuel EJ. Principles for allocation of scarce medical interventions. Lancet 2009;373:423–31

15 Rawls J. A Theory of Justice. Belknap Press, 1971; ISBN 978-0-674-00078-0

16 Nyhan B. Why the “Death Panel” Myth Wouldn’t Die: Misinformation in the Health Care Reform Debate. The Forum, Volume 8, Issue 1;2010 Article 5 The politics of health care reform. Berkeley Electronic Press. www.dartmouth.edu/~nyhan/health-care-misinformation.pdf (accessed 27 July 2020)

17 DIVI (Deutsche Interdisziplinäre Vereinigung für Intensiv-und Notfallmedizin). Entscheidungen über die Zuteilung von Ressourcen in der Notfall-und der Intensivmedizin im Kontext der COVID-19-Pandemie. Klinisch-ethische Empfehlungen. https://www.divi.de/aktuelle-meldungen-intensivmedizin/covid-19-klinisch-ethische-empfehlungen-zur-entscheidung-ueber-die-zuteilung-von-ressourcen-veroeffentlicht (accessed 27 July 2020)

18 Deutsches Ärzteblatt: Zahl der Menschen mit drittem Geschlecht geringer als angenommen. https://www.aerzteblatt.de/nachrichten/102938/Zahl-der-Menschen-mit-drittem-Geschlecht-geringer-als-angenommen (accessed 27 July 2020)

19 Zuckerman P. „Atheism, contemporary numbers and patterns“, in: “The Cambridge Companion to Atheism”, edited by Michael Martin, New York: Cambridge University Press, 2007,47–65

